# 100,000 lumens to treat seasonal affective disorder: A proof of concept RCT of bright, whole-room, all-day (BROAD) light therapy

**DOI:** 10.1101/2021.10.29.21265530

**Authors:** Julia Fabienne Sandkühler, Sarah Brochhagen, Paul Rohde, Rosa Clara Muscheidt, Teja Wolfgang Grömer, Helge Müller, Jan Markus Brauner

**Author notes:** Julia Fabienne Sandkühler (preferred), Abteilung Allgemeine Psychologie I, Institut für Psychologie, Rheinische Friedrich-Wilhelms-Universität Bonn Kaiser-Karl-Ring 9 D-53111 Bonn.

## Abstract

**Background:** Seasonal affective disorder (SAD) is common and debilitating. The standard of care includes light therapy provided by a light box; however, this treatment is restrictive and only moderately effective. Advances in LED technology enable lighting solutions that emit vastly more light than traditional light boxes. Here, we assess the feasibility of BROAD (Bright, whole-ROom, All-Day) light therapy and get a first estimate for its potential effectiveness.

**Methods:** Patients were randomly assigned to a treatment for four weeks; either a very brightly illuminated room in their home for at least six hours per day (BROAD light therapy) or 30 minutes in front of a standard 10,000 lux SAD light box. Feasibility was assessed by monitoring recruitment, adherence, and side effects. SAD symptoms were measured at baseline and after two and four weeks, with the Hamilton Depression Rating Scale-Seasonal Affective Disorders 29-items, self-report version.

**Results:** All 62 patients who started treatment were available at four-week follow-up and no significant adverse effects were reported. SAD symptoms of both groups improved similarly and considerably, in line with previous results. Exploratory analyses indicate that a higher illuminance (lux) is associated with a larger symptom improvement in the BROAD light therapy group.

**Conclusions:** BROAD light therapy is feasible and seems similarly effective as the standard of care while not confining the participants to 30 minutes in front of a light box. In follow-up trials, BROAD light therapy could be modified for increased illuminance, which would likely improve its effectiveness.

## Introduction

Becoming depressed every winter is a common and debilitating experience. Standard light therapy is time-intensive for the patients and only moderately effective. Is the “dose of light” used in the standard therapy simply too low?

Seasonal affective disorder (SAD) is a seasonal pattern of recurrent major depressive episodes that most commonly occurs during autumn or winter with full remission during spring and summer (Zauderer & Ganzer, 2015). SAD is thought to be caused primarily by a lack of daylight (Pjrek et al., 2020); in northern latitudes, the prevalence of SAD is estimated to be up to 10% (Byrne & Brainard, 2008).

Light therapy is one of the most commonly recommended treatments for SAD (Anderson et al., 2008; Association & Others, 2009; Bauer et al., 2013; Kurlansik & Ibay, 2012; Mårtensson et al., 2015; Praschak-Rieder & Willeit, 2003; Ravindran et al., 2009). It is usually provided by sitting in front of a light box for 30 minutes in the morning, restricting the number of activities during this time. This proof of concept trial introduces a new light therapy that enables patients to go about their daily activities during treatment.

Meta-analyses showed that the standard treatment regimen is only moderately effective (Golden et al., 2005; Mårtensson et al., 2015; Pjrek et al., 2020). Since SAD patients remit in summer, the reason why the current treatment is not fully effective might be that it differs too much from summer light. The new light therapy in this trial is more similar to summer than standard light therapy in several ways.

Currently, relevant guidelines and light therapy lamp manufacturers recommend light therapy that has the following characteristics (Kurlansik & Ibay, 2012; MayoClinic, 2017; National Health Service, 2018; National Institute of Mental Health, 20-MH-8138; Philips, 2018):

- Light emitted by a SAD therapy lamp, which usually provides 10,000 lux at 20cm distance to the eyes and only covers a small area of the visual field.
- Exposure for 30 minutes per day, preferably in the morning.

In contrast, consider the characteristics of natural sunlight on a summer’s day:

- *Homogeneous illumination:* The environment is evenly lit, bright light reaches the eye from all of the visual field. It might be beneficial to have the whole visual field illuminated and not just the small area covered by a light box.
- *Duration:* People are exposed to bright illuminance for many hours of the day (Li et al., 2010; Matour et al., 2017).
- *Illuminance:* Sunlight in summer has an illuminance of 9,000 lux to 25,000 lux over the day, in the shadow^1^ (Li et al., 2010; Matour et al., 2017).

Assuming that patients keep the correct distance from the lamps, the standard recommended light therapy broadly meets the illuminance (lux) of natural lighting on a summer’s day. However, it covers only a small part of the visual field, and the duration is short. We aimed to make light therapy more similar to summer lighting by using “more light” - bright light that covers the whole visual field for many hours. We term this experimental treatment BROAD (Bright, whole-ROom, All-Day) light therapy.

Indeed, there are online reports from patients suffering from SAD, who have experimented with such solutions and have reported that they found them much more effective than the standard SAD light box^2^. We interviewed several of these patients, and their experiences directly informed the experimental treatment we evaluated in the trial. The setup for BROAD light therapy, as evaluated in this trial, costs around 350 euros and consists of a socket cord with up to 70 very bright LED light bulbs, which participants installed in a room in their home where they spent at least six hours per day (Figure 1).

**Figure 1.**
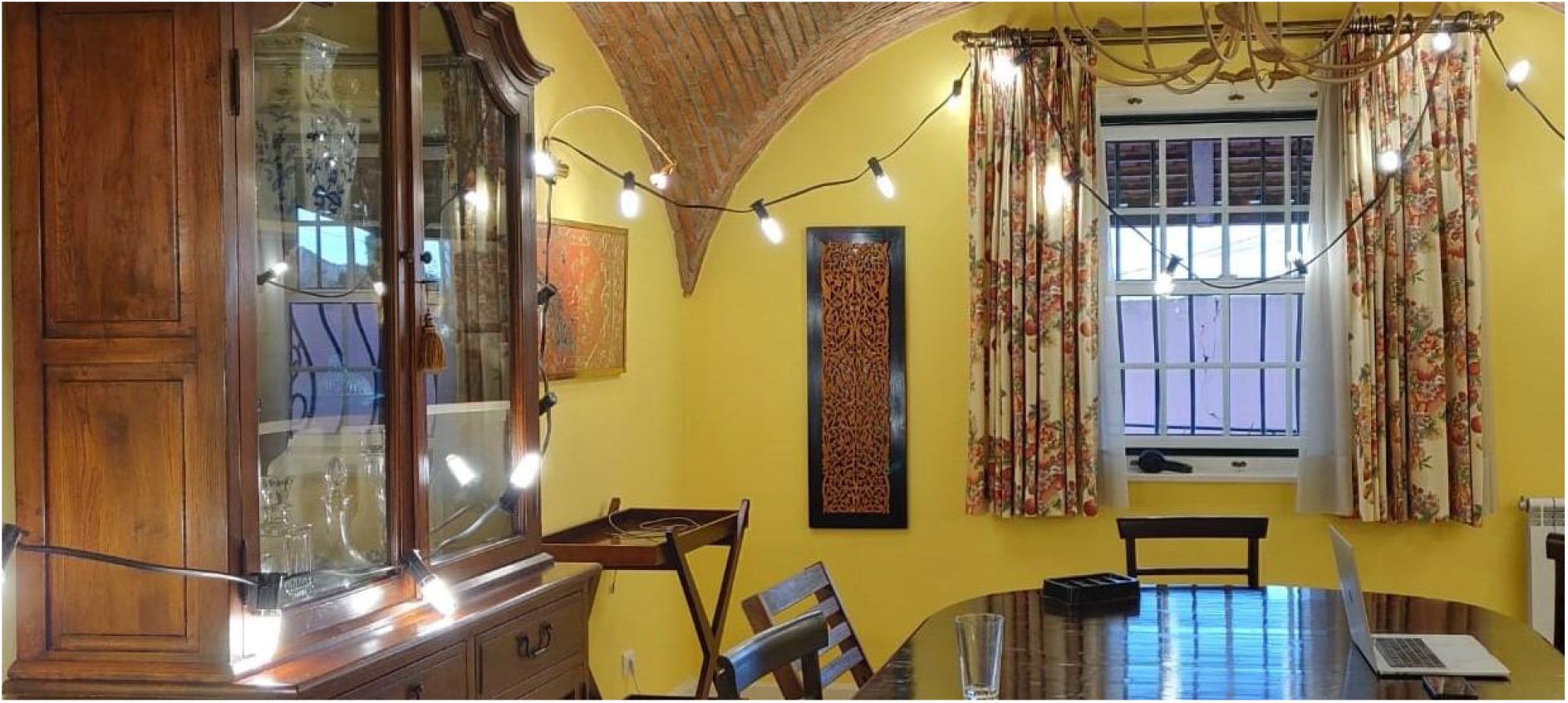
Example BROAD light therapy setup. Participants were provided with one or two such socket cords, a total of 40-70 bright LED light bulbs, and materials to attach them to their room. Only a fraction of the bulbs are shown in this picture. While it is hard to capture on camera, the BROAD setup results in very brightly illuminated rooms.

BROAD light therapy is related to an existing treatment in the literature - light room therapy, i.e. bright light that covers the whole visual field, but which is clinic-based and with shorter treatment durations. Light rooms have been studied for the treatment of SAD, with promising initial results. Four studies on light room therapy against SAD and six studies on light room therapy against other disorders found many positive psychological and neurological effects (Canazei et al., 2017; Kripke et al., 1983, 1992; Rastad et al., 2008, 2011, 2017; Stain-Malmgren et al., 1998; Thalén et al., 1995; Van Someren et al., 1997; Wirz-Justice et al., 1999). These studies gave patients exposure for up to 3 hours with illuminance ranging from 1100 to 4300 lux (Canazei et al., 2017; Kripke et al., 1983, 1992; Rastad et al., 2008, 2011, 2017; Stain-Malmgren et al., 1998; Thalén et al., 1995; Van Someren et al., 1997; Wirz-Justice et al., 1999). No study compared the efficacy of a light room against the more common SAD light box treatment. In all previous studies, the patients had to travel to a central location, usually a clinic, to spend time in a light room specifically installed for this purpose. In contrast, we brightly illuminated a room in the patient’s home for six or more hours per day while they went about their daily activities. This new treatment is cheaper, more convenient for the patients, and enables longer exposure times.

BROAD light therapy seems like a fairly natural extension of light room therapy, so one might ask why it has not been studied before. We think this might be due to past technical constraints. Producing bright light was much harder and more expensive in the past, which made the treatment tested in this trial prohibitively expensive. However, recent advances in LED technology enable affordable and powerful lighting solutions.

Developing less restrictive forms of light therapy is valuable. In addition, we hypothesize that BROAD light therapy can be made more effective at treating SAD than is a SAD light box. Should our hypothesis be confirmed by our proof of concept trial or further follow-up trials, these findings would have the potential to revolutionize SAD therapy and reduce the symptom severity of millions of patients world-wide. The changes to clinical practice would be straight-forward to implement: Physicians could simply recommend different lighting solutions to their patients. In this proof of concept trial, we aimed to explore the feasibility and acceptance of BROAD light therapy, get a first estimate for its potential effect to inform whether larger follow-up trials are warranted, and explore how our concrete implementation of BROAD light therapy can be improved.

## Methods

This is a short summary of the study design and protocol. Further details on all sections can be found in the appendix.

### Study design

We did a randomized, partially blind, parallel-group, proof of concept trial with a primary endpoint at 4 weeks from randomization.The trial evaluated a BROAD light therapy compared with an active control - the standard of care (a SAD light box). The trial design and participant flow are summarized in Figure 2. The trial was prospectively registered (drks.de identifier: DRKS00023075, https://osf.io/ndjm4/) and ethical approval was obtained from the ethics committee of Witten/Herdecke University (184/2020). Apart from telephone calls and the intervention itself, all aspects of the study, including screening, consent, diagnosis video calls, treatment allocation and symptom assessment were conducted online. Participants received the lamps via mail and set them up at home.

**Figure 2.**
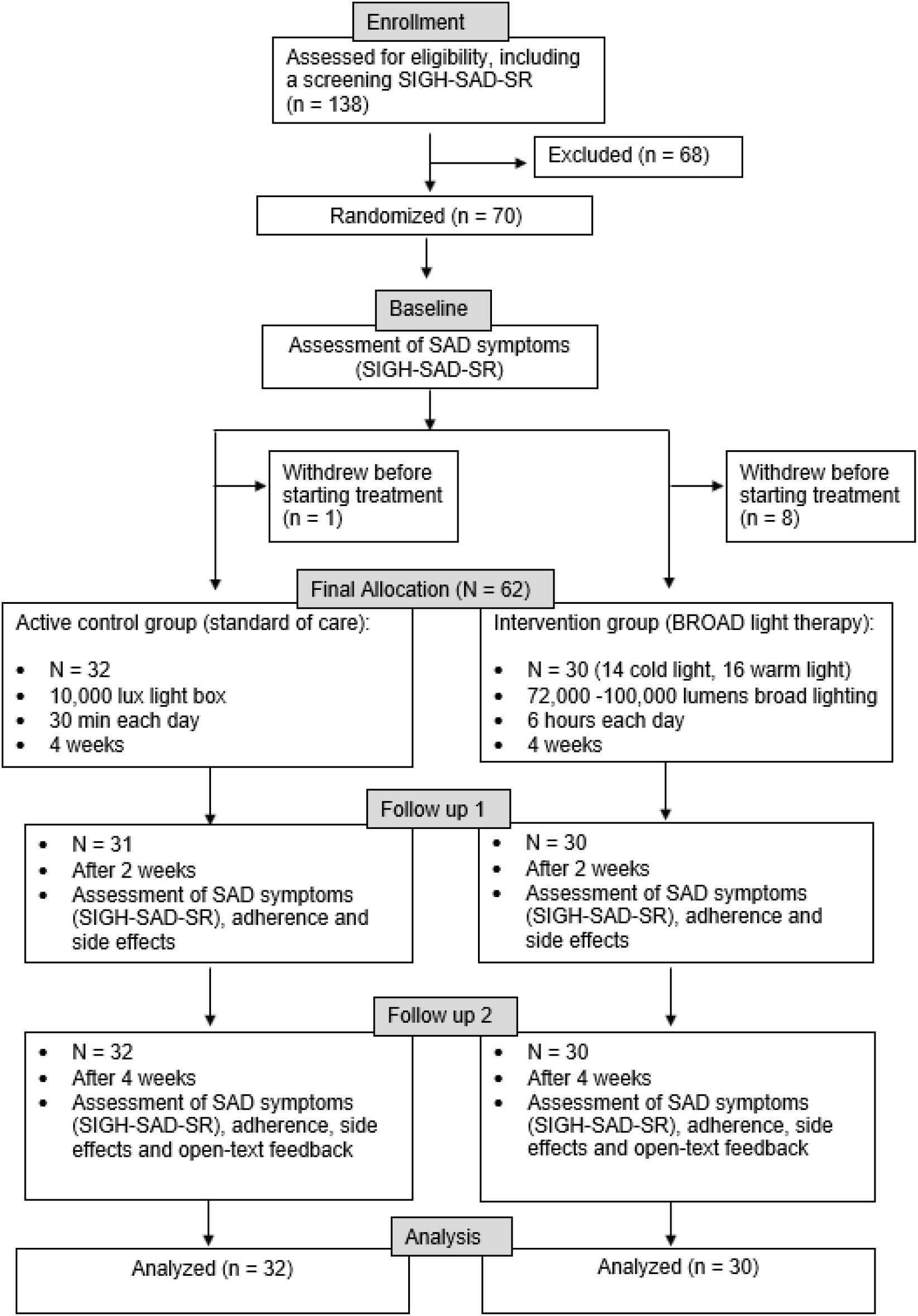
Participant flow through the study.

### Participants

Participants were recruited through general practitioners, clinics, and social media between 29/09/2020 and 29/01/2021 and treated between 17/11/2020 and 17/03/2021. A screening questionnaire briefly assessed if the eligibility criteria were met. Participants who met these criteria received the Hamilton Depression Rating Scale-Seasonal Affective Disorders 29-items Version, self-report version (SIGH-SAD-SR) to assess their SAD symptom severity for screening and randomization purposes. If participants’ symptoms were not yet severe enough, they were asked to answer the screening SIGH-SAD-SR again if they started to feel more depressed.

Participants were 18 years or older with a diagnosis of “major depressive disorder (recurrent episode) with a winter seasonal pattern”, as assessed by a structured clinical interview based on the DSM-V, which was conducted by psychologists in a video call. Patients were eligible to participate in the trial if they were at home for at least 6 hours during morning and afternoon (before 19:00), on at least 5 days per week. Exclusion criteria included a history of manic episodes, light therapy in the previous four months and recent changes in antidepressant medication (see appendix for full list).

At first, the screening information participants received did not adequately convey that they might have to put a long socket cord into their room, which led to some participants in the BROAD light therapy group withdrawing before starting the treatment. After we adjusted the recruitment and screening material, there were no more withdrawals.

### Interventions

#### Active control group

The active control group received a commercially available SAD light box with a color temperature of 6500K and a color rendering index of 80 which provided 10,000 lux at 20cm. With greater distance the illuminance rapidly declined. The lamps were purchased from various manufacturers according to availability and were most commonly the Beurer TL 41 or equivalent lamps. Participants were asked to sit in front of this lamp in the morning at an eye distance of 20cm while e.g. reading or having breakfast. They were asked not to look directly into it apart from occasional squints. The treatment duration was 30 minutes at least five days a week for at least four weeks.

#### Intervention group

There were two BROAD light therapy conditions: a warm condition with a color temperature of 4000K and a cold condition with a color temperature of 6000K. Both had a color rendering index of 80-90. We included these two versions to investigate if there were large differences between them in how acceptable BROAD light therapy is to patients, as warm light is sometimes perceived as more comfortable.

The light in the BROAD light therapy was provided by one or two 10m-20m long socket cords with 20-40 light bulbs each (see Figure 1). The light emitted in the cold condition was 72,000 lumens and 102,240 lumens in the warm condition, so that the amount of blue light was the same in both. This is important because blue light is hypothesised to be the effective wavelength in SAD treatment (Strong et al., 2009). The mean illuminance at eye level when sitting was 1,433 lux (minimum 550 lux, maximum 4,061 lux) in the cold light condition and 1,829 lux (minimum 500 lux, maximum 6,800 lux) in the warm light condition, as measured by the participants with a smartphone app. Despite the BROAD light therapy bulbs emitting vastly more light than the light box (72,000-102,000 lumens vs 850-1,530 lumens), the average amount of light that reached the eye at any given point in time (illuminance) was lower for the BROAD light therapy than the light boxes (1,433-1,829 lux vs 10,000 lux). This was because the light bulbs in the BROAD light therapy setup were much further away from the eyes than the SAD light box (20 cm). The larger distance had the advantage that patients were able to go about their daily activities, enabling a longer treatment duration and more overall illuminance (lux) over time. The treatment duration was at least six hours on at least five days a week for at least four weeks.

#### Similarity of treatment groups

Apart from the inevitable differences in the nature of the treatments, the treatment groups were made as similar as possible, receiving the same questionnaires, checks on the setup and reminders.

### Outcomes

To determine the feasibility of a larger study, we assessed recruitment, adherence, side effects and participant feedback.

To get a first estimate of potential effectiveness, we had one preregistered primary outcome: Difference in SAD symptom severity between baseline and after four weeks of treatment. This outcome was recorded online using the Hamilton Depression Rating Scale-Seasonal Affective Disorders 29-items Version, self-report version (SIGH-SAD-SR) (Terman, Williams, White, Gould, 2008). Our three preregistered secondary outcomes were the difference in SAD symptom severity between baseline and after two weeks of treatment; and the fraction of patients in remission after two and four weeks.

### Randomization and blinding

Patients were randomly assigned (1:1:2) to receive either warm white (4000K) or cold white (6000K) BROAD light therapy or the active control procedure. Randomization was performed using a computer-generated list, in blocks of 4 and 8 participants and in three screening symptom severity strata (SIGH-SAD score <= 19, 20-29, and >= 30) by the senior author, who did not have contact with or knowledge on participants. The senior author received an email with the participant code and symptom severity and emailed back the allocation.

Apart from occasional corrections to the setup of the lamps and answers to participants’ questions, unblinded personnel did not interact with participants. In particular, the outcome surveys were online and included no interaction with the study personnel. Participants did not know which treatment was the active control group nor what all the treatment options were. Participants did not know that there were three groups, or what the difference in treatment between the groups was. However, the lamps provided in the different groups looked different.

### Statistical analysis

The feasibility outcomes were reported descriptively and narratively. In accordance with the CONSORT guideline on pilot and feasibility trials (Eldridge et al., 2016), for the clinical primary and secondary endpoints, we report only descriptive statistics: mean (standard deviation) for continuous outcomes and raw count (%) for categorical outcomes. In an exploratory fashion, we additionally report descriptives of the primary endpoint adjusted for confounders (for details see appendix). In addition, we performed exploratory Pearson correlations between illuminance in the BROAD light therapy condition and the primary endpoint and between measures of adherence and the primary endpoint. The significance level for these tests of correlation was p < .05.

We would have included participants who had dropped out after starting the treatment, but there were none.

## Results

We analyzed all 62 patients who started the treatment. For participant characteristics, see Table 1.

**Table 1.** Baseline characteristics. Data is given as mean (standard deviation) or as number of participants.

|  | Intervention | Active control | Total |
| --- | --- | --- | --- |
| N | 30 | 32 | 62 |
| Age in years (M, SD) | 38.8 (14.5) | 38.5 (15.5) | 38.6 (14.9) |
| Female (N) | 19 | 22 | 41 |
| Male (N) | 11 | 10 | 21 |
| Baseline symptom severity in<br>SIGH-SAD points (M, SD) | 26.3 (6.09) | 27.9 (8.95) | 27.1 (7.68) |

### Feasibility

The recruitment of participants went better than according to plan. The process we had set up to run the study worked and we see no obstacle here for a larger study.

Participants adhered very well to their instructed treatment durations. Both groups underwent treatment for more than the minimum required 5 days a week on average (see Table 2). As instructed, participants in the intervention group underwent treatment for over six hours per day on average, while active control participants underwent treatment for a little over half an hour per day on average (see Table 2).

**Table 2.** Adherence. Data is given as mean (standard deviation).

|  | BROAD light therapy | Light box therapy |
| --- | --- | --- |
| Days treated per week | 5.72 (0.63) | 5.88 (0.79) |
| Hours treated on treatment days | 6.75 (1.28) | 0.6 (0.31) |

No patients discontinued the study due to an adverse event. Some participants (8 in the intervention group and 4 in the active control group) reported side effects of a nature consistent with other studies on light therapy (see Table 3).

**Table 3.** Side effects. Data is given as N (%).

|  | BROAD light therapy | Light box therapy |
| --- | --- | --- |
| Any side effect | 8 (27%) | 4 (13%) |
| Eyeache | 2 (7%) | 2 (6%) |
| Headache | 5 (17%) | 1 (3%) |
| Agitation | 2 (7%) | 1 (3%) |

Qualitative and free-text feedback for both the intervention group and the active control group was generally positive. On a scale from 1 = Completely disagree to 7 = Completely agree, participants in both groups on average somewhat agreed that the treatment had helped them with their symptoms (see Table 4).

**Table 4.** Participant feedback on their treatment. Data is given as mean (standard deviation). Answer options ranged from 1 = Completely disagree to 7 = Completely agree.

|  | BROAD light therapy | Light box therapy |
| --- | --- | --- |
| "I think the light therapy in this study helped me with my symptoms." | 5.33 (1.45) | 5.86 (0.99) |
| "I would recommend this treatment to a friend with similar symptoms to mine." | 5.47 (1.46) | 5.90 (1.23) |

### Effectiveness of BROAD light therapy

This is a proof of concept trial and it is not powered to detect moderate effect size differences between the two treatments. In accordance with the CONSORT guideline on pilot and feasibility trials (Eldridge et al., 2016), we therefore report only descriptive statistics and no tests of significance of the differences.

Our primary endpoint, the improvement in symptoms after four weeks of treatment, was considerable and similar for all treatment groups (see Table 5 and Figure 3). The magnitude of SAD symptom improvement after light therapy is in line with previous results (Pjrek et al., 2020). The same was true for our secondary endpoints, symptom improvement after two weeks of treatment and remission (defined as a SIGH-SAD score below 9) at two and four weeks (see Table 5). The improvement in symptoms was similar for the cold and the warm color temperature BROAD therapy groups (see appendix).

**Table 5.** Secondary endpoints. Mean (standard deviation) [95% confidence intervals of the mean improvement] in SIGH-SAD points given for unadjusted symptom improvement and N (%) given for remission. The adjusted symptom improvement does not correspond to SIGH-SAD points, but only serves as a unitless measure of similarity.

|  | BROAD light therapy | Light box therapy |
| --- | --- | --- |
| Symptom improvement after 4 weeks | 11.58 (8.39), [8.45; 14.72] | 13.24 (9.64), [9.76; 16.72] |
| Symptom improvement after 4 weeks, adjusted (arbitrary units) | 12.56 (7.66), [9.70; 15.42] | 12.92 (9.19), [9.61; 16.23] |
| Subscale A symptom improvement after 4 weeks | 7.22 (5.32), [5.23; 9.20] | 8.21 (6.55), [5.85; 10.57] |
| Subscale B symptom improvement after 4 weeks | 4.37 (4.49), [2.69; 6.04] | 5.03 (4.32), [3.47; 6.59] |
| Symptom improvement after 2 weeks | 8.92 (8.75), [5.65; 12.18] | 10.8 (8.16), [7.80; 13.79] |
| Remission after 2 weeks | 3 (10%) | 3 (10%) |
| Remission after 4 weeks | 7 (22%) | 6 (20%) |

**Figure 3.**
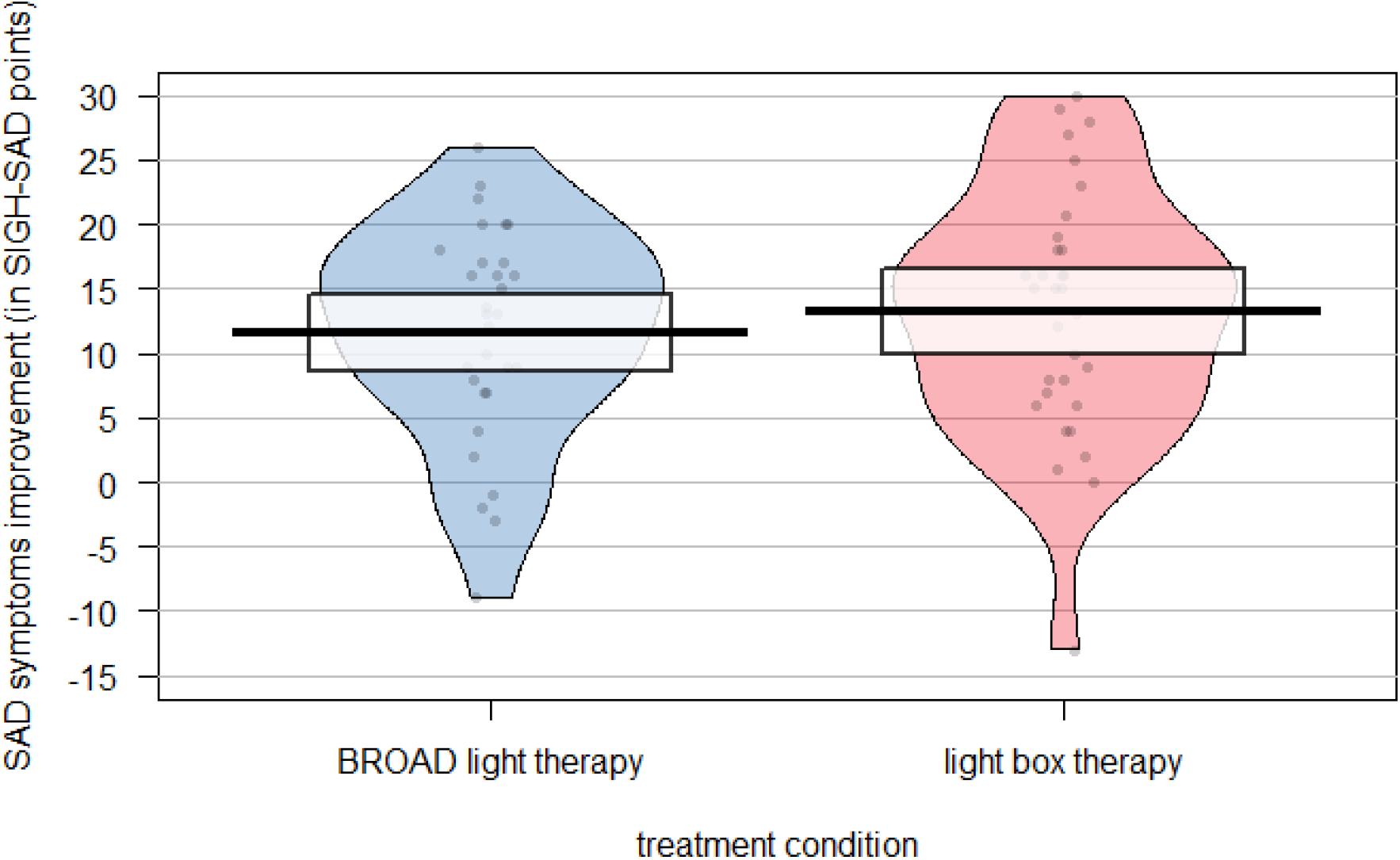
Improvement in SIGH-SAD score after four weeks of treatment. The plot shows the means, 95% confidence intervals of the means and distribution for both the light box group and the BROAD group.

When adjusting for baseline symptom severity, timepoint of treatment and delay between baseline assessment and start of treatment, the improvement after four weeks was even more similar between treatments (see Table 5 and the appendix). The SIGH-SAD consists of two subscales, measuring typical and atypical symptoms of major depression respectively. The improvements in both scales were very similar for both treatments.

### More light, better results

In the intervention group, the illuminance (lux) at eye level depended on the room sizes and how/where the light bulbs were attached. Accordingly, the illuminance at eye level varied greatly between participants in the BROAD light therapy group (between 500 lux and 6800 lux). The illuminance was lower than for the active control group under ideal conditions, because the BROAD light therapy bulbs were further away from the eyes (see Methods - Interventions). This allowed participants to go about their daily activities and thus enabled a longer treatment duration per day and more total lux. The higher the illuminance in the BROAD light therapy group, the more participants’ symptoms improved after four weeks, *r*(28) = 0.43, *p* = 0.032 (see Table 6). As can be seen in Figure 4, every doubling of illuminance (lux) in the intervention group predicted an increase in symptom improvement by 4 SIGH-SAD points on average. To further display this trend, we can split the participants that received BROAD light therapy into two groups: Participants who received above median illuminance at eye levels (>1400 lux) experienced a mean improvement of 16.7 points on the SIGH-SAD score (SD = 6.5), which is more than in the active control group (which improved by M = 13.24, SD = 9.64), while the participants with below median illuminance improved by only 8.9 points on average (SD = 8.5). In addition, treatment hours per day correlated positively and bordering on significance with symptom improvement for the BROAD light therapy group. However, days treated per week did not (see Table 6).

**Table 6.** Correlations with symptom improvement four weeks after the start of treatment for the BROAD light therapy group. * significant at *p* < .05.

|  | Pearson's r | p-value |
| --- | --- | --- |
| Log<sub>2</sub>(lux) and symptom improvement | 0.43 | 0.032* |
| Hours treated per day and symptom improvement | 0.34 | 0.070 |
| Days treated per week and symptom improvement | 0.05 | 0.801 |

**Figure 4.**
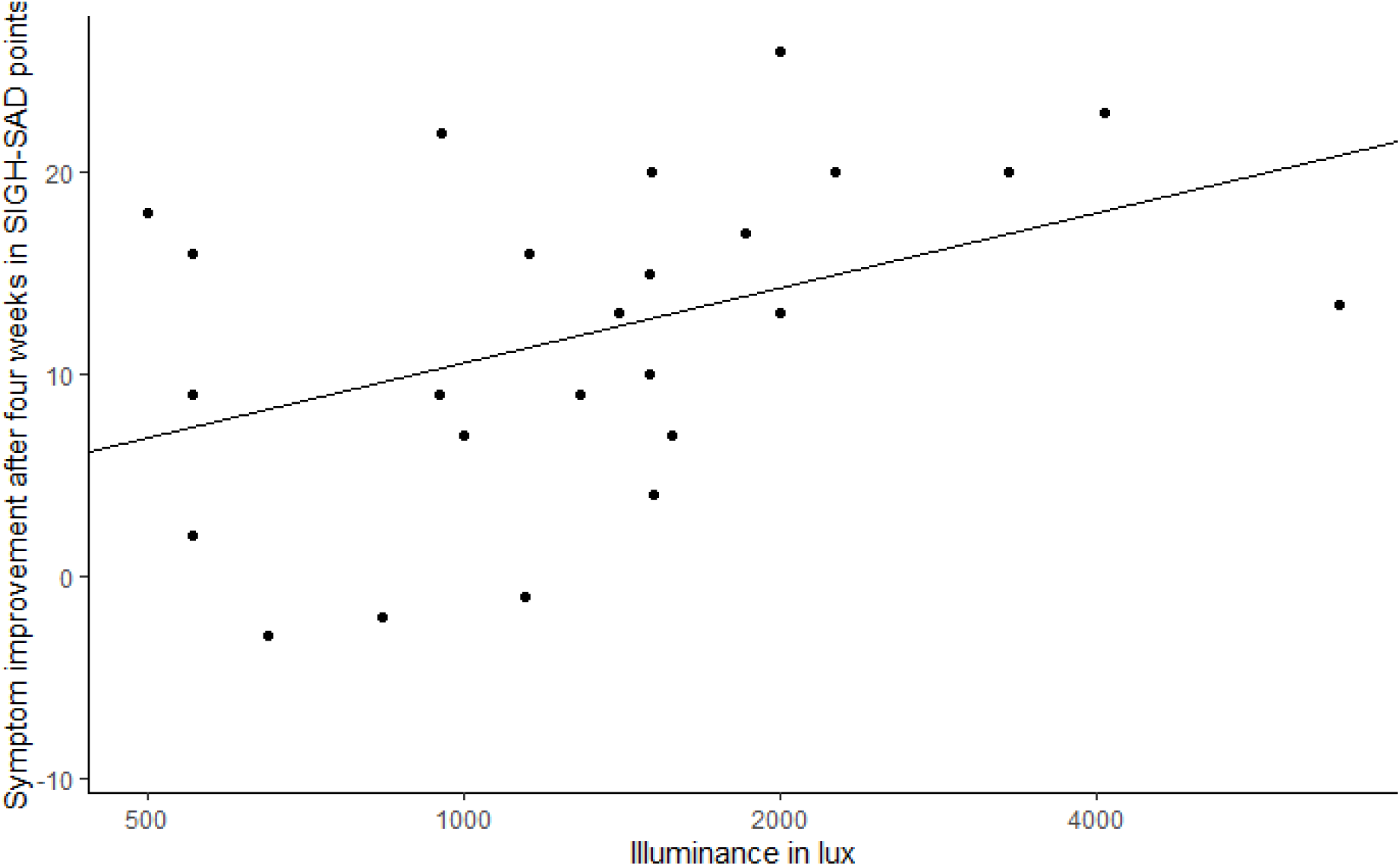
Correlation between symptom improvement after four weeks and the illuminance in the BROAD light therapy group in lux. Participants who received more lux had better outcomes. Scatter plot with trendline.

## Discussion

This is the first study on the feasibility of BROAD light therapy (Bright, whole-ROom, All-Day light therapy). This is also the direct first comparison of any light-room treatment (whether clinic-based or at home, for 2 hours or all day) with the current standard of care, a SAD light box.

In this proof of concept trial, treatments were roughly equally effective on average. The improvement in symptom severity (measured by the SIGH-SAD-SR score) in our study was similar to those of other light therapy studies (Pjrek et al., 2020). Could non-inferiority of BROAD light therapy treatment to standard of care be confirmed in a larger trial, this would be an exciting result for patients. To receive 10,000 lux, patients need to sit very closely to the light box (distance from eyes to light box of 20 cm are common for commercial light boxes), meaning there is half an hour each morning in which they can do little else. BROAD light therapy puts no such constraint on patients’ daily activities, possibly improving long-term adherence. To illustrate: In the course of the four weeks of therapy in this study, control participants spent 14 hours in front of the light box. For the same reason, BROAD light therapy could be a good candidate for prevention, which many patients are likely to only do if it is very low effort. Additionally, we carefully instructed participants on how to use the light box to receive 10,000 lux - in non-study conditions, many participants will likely position themselves further away from the light box than 20 cm, thus receiving a lower illuminance. Again, there is no analogous concern for BROAD light therapy.

Furthermore, we have reason to believe that the BROAD light therapy treatment in this study can be modified to improve its effectiveness. In the BROAD light therapy group, the illuminance (lux) at eye level depended on the room sizes and how/where the lights were attached. Accordingly, the illuminance at eye level varied greatly between participants in this group (between 500 lux and 6800 lux). Exploratory analyses found an interesting significant correlation: The more light reached the participants’ eyes, the more participants’ symptoms improved after four weeks. Participants who received above median illuminance at eye levels experienced a larger improvement than in the active control group, while the participants with below median illuminance improved by less than the active control group. Additionally, undergoing BROAD light therapy for more hours per day was borderline significantly correlated with treatment success - however, here the causation may be the other way around, with participants who experienced the treatment to be more effective using it more regularly or for longer periods. These results offer some support for the idea behind our study, that “more light” leads to better results for patients.

## Limitations

As expected, the variance in our proof of concept trial was too large given our sample size to give much indication of whether one of the treatments performs better than the others. Additionally, illuminance levels in the BROAD light therapy group were likely too low for some participants, so that this treatment was not as effective as it could be.

In addition, the light boxes were marketed on their packaging for the use against SAD, while the components of the BROAD light therapy were not, possibly leading to a higher placebo effect in the active control group than in the BROAD light therapy group. In a follow-up study, this limitation can be addressed by unifying the packaging that lamps arrive in.

Lastly, we did not include a placebo or waitlist control group, so we cannot be certain that the improvement in our participants was due to the treatments. However, the effectiveness of the active control treatment has been well-studied and the size of the improvement in our study are consistent with previous findings.

## Conclusion

It is plausible that increasing the amount of light in light therapy makes it more effective against SAD. Our proof of concept trial did not find BROAD light therapy to be superior to light boxes, but it adds some first empirical evidence that BROAD light therapy is feasible and that the amount of light that reaches participants’ eyes (illuminance/lux) predicts how effective it is at treating SAD. There are several straightforward ways to further increase the brightness of the treatment studied in this paper, potentially creating a treatment that is superior to the current standard of care.

## Data Availability

All data produced are available online at https://osf.io/ndjm4/.

## Acknowledgements

We are grateful to Eliezer Yudkowsky, as we took the inspiration for this trial from his book “Inadequate Equilibria”, and he helped with raising funding and contributed ideas to the initial study design. We thank Johann-Lukas Voigt and Simon Grosse-Holz for help with calculating the fraction of blue light in LEDs of various color temperatures. We are grateful to Prof. Ulrich Ettinger for his feedback and support.

## Data

The data of this study are openly available at the Open Science Framework, https://osf.io/ndjm4/.

## Footnotes

1 Due to their position in the eye socket, the eye balls are usually in the shadow. On surfaces with direct sun exposure, much higher illuminances of more than 100,000 lux are reached during the summer (Li et al., 2010; Matour et al., 2017).

2 Example from a SAD patient: https://meaningness.com/metablog/sad-light-led-lux. Example from a healthy person: https://www.benkuhn.net/lux/

