## Appendix for "100,000 lumens to treat seasonal affective disorder: A proof of concept RCT of bright, whole-room, all-day (BROAD) light therapy" (from another version of this preprint: version 1, 2021-10-30)

**Methods** **2**
1.1. Details of recruitment and procedure 2
1.2. Full list of criteria for inclusion and exclusion 2
1.3. Randomisation and masking 3
1.4. Details on the active control treatment 4
1.5. Details on the BROAD light therapy treatment 4
1.5. Details on statistical analysis 4
1.6. Sample size calculation 4
**Results** **5**
2.1. Why the trial ended or was stopped 5
2.2. Details of withdrawals 5
2.3. Cold and warm BROAD light therapy subgroups 5
2.4. Adjustment 6
2.5. Correlation graphs: Symptom improvement and adherence 8
**References** **9**

### 1. Methods

#### 1.1. Details of recruitment and procedure

Participants were recruited through general practitioners, clinics, and social media. Participants were provided with an information sheet for the study and a consent form for the screening. The information sheet made it clear that they would be randomly assigned to receive different forms of light therapy at home. If consent was given, the participant filled out an online screening form. Demographic information including age and sex as well as questions screening for SAD and for being able to take part in the BROAD light therapy condition were obtained from the participants at screening. The SIGH-SAD-SR was used to assess severity and intensity of recent SAD symptoms at screening and as the basis for randomization strata. The SIGH-SAD-SR has been previously shown to be reliable and valid (1).

Participants who passed the screening were given a structured clinical interview of the DSM-V by a psychologist from the hospital in Herdecke to determine if they had major depressive disorder (recurrent) with a winter seasonal pattern. The interview was conducted over Skype and took about 30 minutes. It included a diagnosis of an episode of major depression, a seasonal pattern and a differentiation from bipolar disorder and dysthymia. Participants who received the diagnosis filled out a consent form for the study. Participants had the opportunity to ask questions concerning the study. If consent was given, the participant was enrolled into the study. The SIGH-SAD-SR was completed again. This up-to-date assessment of a participant's SAD symptoms was used as the pre-treatment baseline score.

Participants were then randomly assigned into the treatments and informed of allocation status. They received instructions on how to set up and undergo their allocated treatment and received their treatment components via post (daylight lamp for the active control group or socket cord, bulbs and adhesive hooks for the BROAD light therapy group). Participants sent a photo of their treatment setup, and in the case of the BROAD light therapy, a lux measurement at eye level using a mobile phone app and received feedback on the setup. If the setup was adequate, participants started the treatment.

#### 1.2. Full list of criteria for inclusion and exclusion

Criteria for inclusion:

- patients suffering from "major depressive disorder (recurrent) with a winter seasonal pattern" as diagnosed by a structured clinical interview of the DSM-V
- at home for at least 6 hours during morning and afternoon (before 19:00), on at least 5 days per week

Criteria for exclusion:

- patients who have used light therapy in the previous four months to avoid carryover effects

- patients with ocular diseases or who are taking photosensitising medication to reduce the risk of damaging their eyes,
- patients who do shift work, because we want to study the effects of light therapy in the morning after a night of sleep, which is not possible for these people,
- pregnant participants, in case this new treatment has any unforeseen negative effects on unborn babies,
- patients who have started or stopped taking melatonin in the previous four months, because this is a different treatment for winter depression that might introduce too much variance,
- patients who have started taking a new antidepressant in the previous six weeks, because this would introduce too much variance,
- patients who are not fit to consent,
- patients with a history of at least one manic episode in the last ten years

#### 1.3. Randomisation and masking

Patients were randomly assigned (1:1:2) to receive either warm white (4000K) or cold white (6000K) BROAD light therapy or the active control procedure (standard of care - commercially available daylight therapy lamps). We decided on this allocation ratio because our main interest lay in comparing the BROAD light therapy with the active control. We did not expect there to be any difference in effectiveness between the two different BROAD light therapy groups.

Diagnosis interviewers regularly sent participant codes and baseline symptom severity (SIGH-SAD) scores of patients who were ready for randomisation to the senior author in charge of randomization, who was not involved in recruitment and did not know which patients were being randomised. He randomised participants and sent allocations back to the research team. Randomisation was done with Excel in blocks of 4 or 8 participants and based on symptom severity (baseline SIGH-SAD score) strata (<= 19, 20-29, >= 30) to ensure that the two trial groups were not significantly different on baseline symptom severity. The block sizes and SIGH-SAD score strata were unknown to anyone but the senior author.

Apart from the structured interview for diagnosis and eligibility, which happened before randomisation, interaction between personnel and patients was minimal. Interactions during which personnel were unblinded consisted of standardised emails and SMS/Whatsapp messages reminding patients to answer the questionnaires or standardised feedback on the setup of the daylight lamp or the BROAD light therapy. On some occasions, we needed to communicate with patients in a less standardised way to help them improve the setup or to answer patients' questions. There was no interaction between personnel and patients at data collection, as patients filled out the questionnaires online from home.

Participants did not know which treatment was the active control group nor what all the treatment options were. Participants did not know that there were three groups, or what the difference in treatment between the groups was. However, the lamps provided in the different groups looked different. Participants knew that there was the possibility that they would have to hang up a cord with light bulbs in their workspace, which might involve adhesive hooks on their wallpaper.

#### 1.4. Details on the active control treatment

The SAD lamps used for the active control group were the Beurer TL 41, Beurer TL 40 and Toplife TL 1880. The only difference between the two Beurer lamps is that one has a switch and the other a touch button. The Toplife lamp is smaller and flatter than the Beurer lamps. They give the same light intensity at 20cm distance from the eyes but the Toplife lamp illuminates a smaller area.

#### 1.5. Details on the BROAD light therapy treatment

Natural daylight has these color temperatures at different times throughout the day, the warmest color temperature occurring at sunset/sunrise at about 3200K and the coldest at noon at about 7200K (2).

#### 1.5. Details on statistical analysis

The statistical analysis was performed with R.

#### 1.6. Sample size calculation

We preregistered (https://osf.io/ndjm4/) that we would allocate 176 participants to treatments in a larger trial, but would do an intermediate analysis and potentially change the trial design based on the results. In the first winter (2020/21), we enrolled 62 participants, because this is what we could comfortably do in one winter. As mentioned in the preregistration, we are herewith publishing these intermediate results as a proof of concept trial and will adjust the trial design based on these results.

### 2. Results

#### 2.1. Why the trial ended or was stopped

Recruitment stopped when new participants could not be expected to be included in time to complete four weeks of treatment before the winter ended.

#### 2.2. Details of withdrawals

Overall, 70 participants were randomized to double-blind treatment; of these, 8 dropped out before the start of the treatment (one in the active control group, four in the cold BROAD light therapy  group, three in the warm BROAD light therapy group). We did not include participants in our analysis who had been randomised but who withdrew before starting the treatment. Early in the study, the participant information was not clear enough on the possibility that participants would have to install a long and heavy socket cord, which led to some dropouts in the BROAD light therapy group before the start of the treatment. We adjusted the screening survey in response and subsequently there were no dropouts anymore.

All but one of the participants who started the treatment completed the full course of treatment. One active control participant completed only the first 12 days of treatment because of time constraints and missed the SIGH-SAD questionnaire two weeks after start of treatment but was again available for the SIGH-SAD questionnaire four weeks after start of treatment.

#### 2.3. Cold and warm BROAD light therapy subgroups

| | Warm BROAD light therapy | Cold BROAD light therapy |
|---|---|---|
| N | 16 | 14 |
| Symptom improvement after 4 weeks | 12.22 (8.40), [7.74; 16.70] | 10.86 (8.64), [5.87; 15.84] |
| Symptom improvement after 2 weeks | 7.34 (7.72), [3.23; 11.46] | 10.71 (9.77), [5.07; 16.36] |
| Remission after 2 weeks | 0 (0%) | 3 (21.4%) |
| Remission after 4 weeks | 2 (12.5%) | 4 (28.6%) |

**Table 1.** Means and standard deviations of the symptom improvement (in SIGH-SAD points) for the two subgroups of the BROAD light therapy group.

#### 2.4. Adjustment

The adjustment of the primary outcome, symptom improvement after four weeks, was based on a regression with improvement as the predicted variable and treatment (active control, intervention), baseline symptom severity, timepoint of treatment and days between baseline assessment and treatment (delay) as predictors.

We took a number of measures to reduce the differences in these three possible confounders between treatment groups. We randomized participants in blocks of 4 and 8 participants and in three screening symptom severity strata (SIGH-SAD score <= 19, 20-29, and >= 30). The light box was sent with a purposeful delay after baseline assessment to participants in the light box therapy group to match the delay in the BROAD light therapy group, which was due to the longer shipping duration and time needed to set up the treatment. Despite these measures, there were slight differences between the two treatment groups (Table 2).

| | BROAD light therapy | Light box therapy |
|---|---|---|
| Baseline symptom severity [SIGH-SAD points] | 26.28 (6.09) | 27.9 (8.95) |
| Timepoint of treatment start | 17/01/2021 [29/11/20 - 01/03/21] | 12/01/2021 [17/11/20 - 17/02/21] |
| Days between baseline and start of treatment (delay) | 16.63 (7.47) | 13.38 (7.53) |

**Table 2.** Data is given as mean (standard deviation) or as median [range].

An adjusted score for symptom improvement was calculated by taking the original symptom improvement score and deducting from it: 1) the baseline score multiplied by the baseline coefficient (0.75), 2) the timepoint of treatment multiplied by the timepoint coefficient in system time (1.06e-06), and 3) the delay multiplied by the delay coefficient (-0.17).

We adjusted for these three confounders because they seem to us to be theoretically plausible influences on symptom improvement and because they correlated in the same direction with symptom improvement in both treatment groups (Figures 1-6).

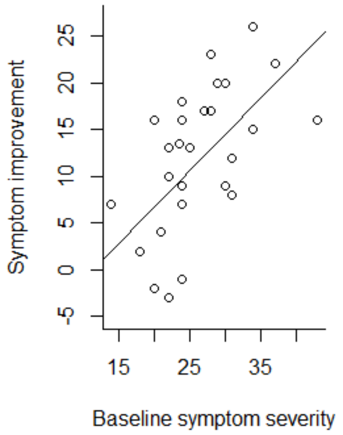


**Figure 1.** Correlation between symptom improvement after four weeks in SIGH-SAD points and baseline symptom severity in SIGH-SAD points for the BROAD light therapy group.

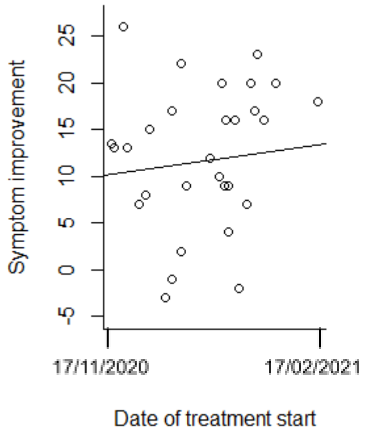


**Figure 2.** Correlation between symptom improvement after four weeks in SIGH-SAD points and date of treatment start for the BROAD light therapy group.

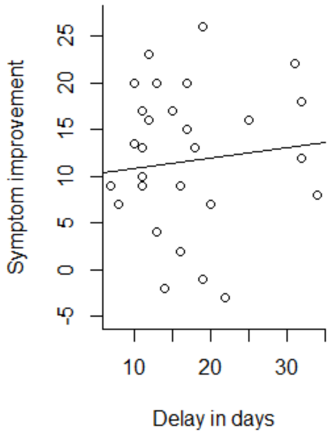


**Figure 3.** Correlation between symptom improvement after four weeks in SIGH-SAD points and days between baseline and start of treatment for the BROAD light therapy group.

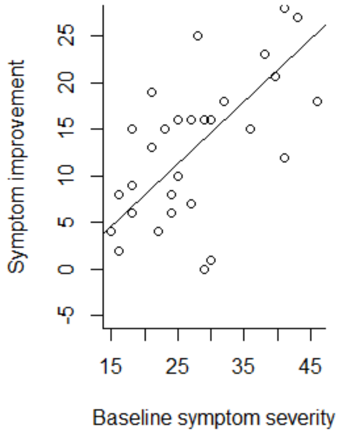


**Figure 4.** Correlation between symptom improvement after four weeks in SIGH-SAD points and baseline symptom severity in SIGH-SAD points for the light box therapy group.

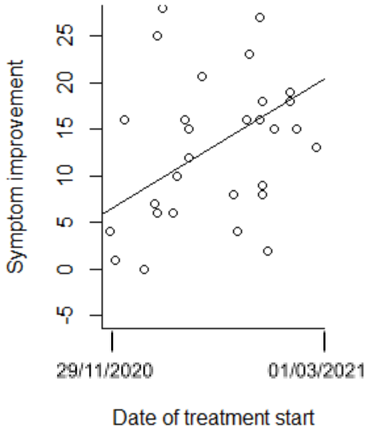


**Figure 5.** Correlation between symptom improvement after four weeks in SIGH-SAD points and date of treatment start for the light box therapy group.

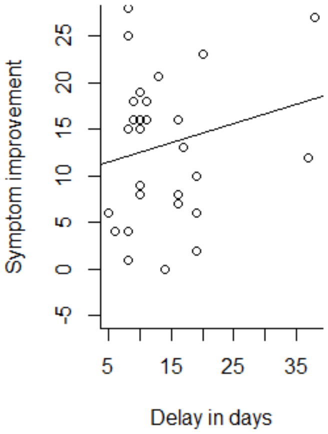


**Figure 6.** Correlation between symptom improvement after four weeks in SIGH-SAD points and days between baseline and start of treatment for the light box therapy group.

#### 2.5. Correlation graphs: Symptom improvement and adherence

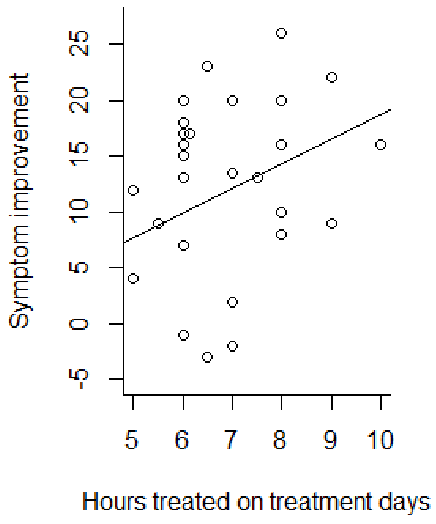


**Figure 7.** Correlation between symptom improvement after four weeks in SIGH-SAD points and hours treated on treated days for the BROAD light therapy group.

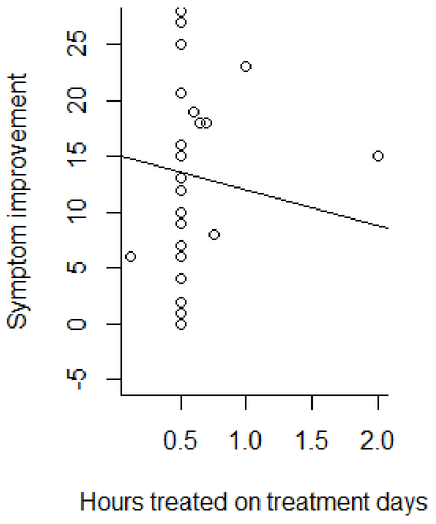


**Figure 8.** Correlation between symptom improvement after four weeks in SIGH-SAD points and hours treated on treated days for the light box therapy group.

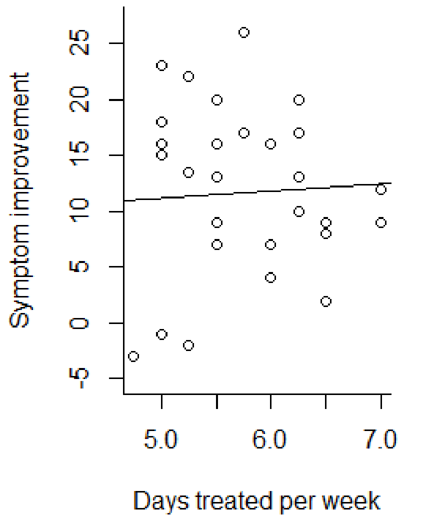


**Figure 9.** Correlation between symptom improvement after four weeks in SIGH-SAD points and days treated per week for the BROAD light therapy group.

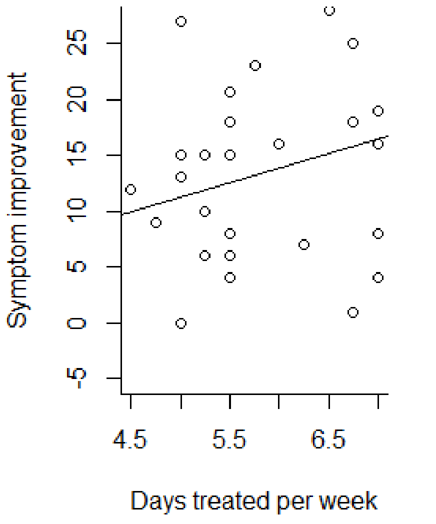


**Figure 10.** Correlation between symptom improvement after four weeks in SIGH-SAD points and days treated per week for the light box therapy group.
